# Neighborhood Disadvantage Association with Sleep Apnea and Longitudinal Cardiovascular Events in a Large Clinical Cohort

**DOI:** 10.1101/2024.01.31.24302108

**Authors:** Cinthya Pena-Orbea, David Bruckman, Jarrod E. Dalton, J. Daryl Thornton, Jay L. Alberts, Catherine M. Heinzinger, Nancy Foldvary-Schaefer, Reena Mehra

**Affiliations:** Sleep Disorders Center, Neurological Institute, Cleveland Clinic, Cleveland OH; Center for Populations Health Research, Department of Quantitative Health Sciences, Lerner Research Institute, Cleveland Clinic, Cleveland, OH; Center for Health Equity Engagement, Education and Research, The MetroHealth Campus of Case Western Reserve University, Cleveland, OH; Population Health and Equity Research Institute, The MetroHealth Campus of Case Western Reserve University, Cleveland, OH; Division of Pulmonary, Critical Care, and Sleep Medicine, The MetroHealth Campus of Case Western Reserve University, Cleveland, OH; Department of Biomedical Engineering, Cleveland Clinic, Cleveland, Ohio; Center for Neurological Restoration, Cleveland Clinic, Cleveland, Ohio; Respiratory Institute, Heart and Vascular Institute and Lerner Research Institute, Cleveland Clinic, Cleveland

## Abstract

**Background:** The association between neighborhood socioeconomic disadvantage and poor cardiovascular outcomes is well established; however, less is known about its interplay with obstructive sleep apnea.

**Methods:** Adult cardiovascular disease-naïve patients who underwent sleep testing at Cleveland Clinic in Ohio from August of 1998 to August of 2021 were included in this cohort. The primary exposure was Area Deprivation Index (ADI) calculated by national rank, i.e. 25^th^, 50^th,^ and 75^th^ percentiles; higher quartiles reflecting greater deprivation (ADI-Q1-4) with Q1 as reference. Cox proportional hazard models were used to determine the hazard of composite outcome of major adverse cardiovascular events (MACE), i.e. including heart failure, stroke, atrial fibrillation and coronary artery disease or death, adjusted for demographics, comorbidities, cardiac medications and objective OSA-related measures including of Apnea Hypopnea Index (AHI) and sleep-related hypoxia (percentage of sleep time spent<90%SaO2,T90). Linear models were used to examine the relationship between ADI and OSA-related measures. Interaction terms were tested between ADI and OSA-related measures.

**Results:** Of 72,443 adults age was 50.4±14.2 years, 50.5% were men, and 18.4% Black individuals. The median AHI was 14.3[5.8, 33.3] with a median follow-up of 4.39 [IQR,1.76-7.92] years. The relative incidence of initial MACE in the presence of competing risk of death was 17% higher (HR,1.17[95%CI 1.09-1.27],p<.001) for those living in ADI-Q4. Greater levels of area deprivation were associated with sleep-related hypoxia measures including higher degree of T90(p<.001); lower mean SaO2(p<.001), and lower minimum SaO2(p<.001). Significant interactions between T90 and ADI were observed with the risk of MACE(p=0.002) or death(p=0.005). T90 conferred a 37% increased risk of MACE(HR, 1.37[95%CI:1.23-1.53]) for those living in ADI-Q1; and a 26% increased risk(HR, 1.26[95%CI:1.14-1.38%]) among patients living in ADI-Q4. For individuals living in ADI-Q2 and Q3, T90 conferred a respective 56% and 51% increased risk of death (HR,1.56[95%CI:1.23 - 1.96]; HR, 1.51[95%CI:1.21-1.88]), respectively.

**Conclusions:** Neighborhood disadvantage was associated with an increased risk for MACE or death in this clinical cohort and this association was modified by sleep-related hypoxia. Further research is needed to identify neighborhood-specific social determinants contributing to sleep-cardiovascular health disparities to develop neighborhood-specific interventions.

**Clinical Perspective:** *What is new?:*

- This is the largest-to-date longitudinal study using a large clinically phenotyped sample that uncovers the association of neighborhood socioeconomic position and sleep apnea with major cardiovascular events and mortality.
- In this cohort, patients with increased sleep-related hypoxia living in both extremes of area deprivation had an increased risk for major adverse cardiovascular events, whereas individuals with increased sleep-related hypoxia living in moderate areas of deprivation had an increased risk for death.

*What Are the Clinical Implications?:*

Addressing disparities in sleep-related hypoxia may be a modifiable and targetable intervention to decreased overall health disparities cardiovascular health.
There is a call to action to develop future studies examining neighborhood-level social determinants of health that influence increased sleep-related hypoxia in patients with sleep apnea to improve cardiovascular outcomes across all populations at risk for health inequities.

## Introduction

Despite substantial progress in reducing cardiovascular disease (CVD)-related morbidity and mortality, CVD remains the leading cause of death in the United States.^1,2^ Over the past decade, CVD prevention and treatment have markedly advanced, however, disparities in CVD prevalence, risk factors, and health outcomes across different racial and ethnic and socioeconomic position (SEP) groups persist.^3–6^ Evidence has consistently shown that socioeconomic deprivation is an important and underrecognized determinant of cardiovascular health accounting for 44% of geographic disparities in cardiovascular mortality.^7^ For instance, in ischemic heart disease, low SEP has been linked with increased disease burden, less access to care, and higher mortality rates.^8,9^ Furthermore, residence in socioeconomically disadvantaged communities has been associated with increased risk of rehospitalization and death in patients with heart failure and myocardial ischemia after accounting for an individual’s SEP factors ^10^ Therefore, understanding factors that contribute to SEP-related disparities in CVD is needed to inform equitable approaches to improve cardiovascular health.

Obstructive Sleep Apnea (OSA) is a highly prevalent condition that affects 40-80% of individuals with CVD and is associated with an increased prevalence of cardiovascular (CV) risk factors and CVD-related morbidity and mortality. As such, sleep disturbances have become recognized as a target to improve CV health and a component of the American Heart Association Life’s Essential 8.^11,12^ Emerging data identify racial and ethnic disparities in OSA severity, diagnosis, and treatment.^13^ ^14^ Moreover a few studies in the pediatric population have identified SEP to be associated with increased OSA severity. ^15–18^ However, the influence of OSA as a potential contributor to socioeconomic disparities in CV health outcomes has received minimal attention.

The area deprivation index (ADI) is a validated neighborhood marker of socioeconomic disadvantage consisting of a range of socioeconomic indicators including poverty, education, housing, and employment.^19^ Several studies have shown that living in areas with high ADI is associated with an increased incidence of CVD and worse cardiovascular outcomes.^7,20–22^ Although the association between ADI and increased CV outcomes is well-established, a critical existing knowledge gap is lack of understanding of the association of area deprivation and CV outcomes in OSA and how sleep disturbances may influence socioeconomic disadvantage in relation to CV outcomes. We hypothesize that living in greater areas of disadvantage is associated with an increased incidence of major adverse cardiovascular events (MACE) and mortality in a clinical sleep referral cohort and that these associations will be modified by the degree of OSA severity.

## Methods

A retrospective cohort study was conducted using the STARLIT Registry (Sleep Signals, Testing, and Reports Linked to Patient Traits) at the Cleveland Clinic. First, we investigated the association between ADI and a composite incident MACE outcome and all-cause mortality in a large cohort of patients evaluated for sleep disorders. Second, we examined the association of objective measures of ADI and OSA. Finally, effect modification of OSA on the association of area deprivation and CV outcomes was assessed. The study was approved by the Cleveland Clinic IRB as a minimal-risk research study for which informed consent was waived. This study followed the Strengthening the Reporting of Observational Studies in Epidemiology (STROBE)^23^ reporting guideline for reporting of cohort studies.

### Study Populations and Data Source

We identified patients from the STARLIT Registry **(see Supplemental Methods for registry details)** without established CVD (any history of atrial fibrillation, heart failure, cerebrovascular events, and coronary artery disease) at the time the sleep study was performed and who subsequently underwent follow-up. Patients were included if they were > 18 years old, had a diagnostic sleep study [polysomnogram (PSG)], split PSG, or type III sleep study with a minimum of ≥3 h diagnostic time available, naïve to OSA treatment, a resident of Ohio at or before the sleep study and if they lived in an identifiable census tract. Demographics, primary payor, body mass index kg/m^2^ (BMI), comorbidities, cardiovascular medications, and objective OSA-related measures were extracted from the sleep study registry and medical record (**see Table S1 for cardiovascular medications and comorbidities details**). The latest version of the Elixhauser Comorbidity Index (V2021.1)^24^ was used to evaluate for medical complexity and utilized due to the optimal performance in cardiac conditions.^25^ All sleep studies were conducted, and respiratory events were scored in accordance with the American Academy of Sleep Medicine guidelines.^26^ Objective OSA-related measures of interest included frequency of apneas and hypopneas (apnea hypopnea index, AHI) and sleep-related hypoxemia (percentage of sleep time spent ≤90%SaO2 [T90]). The latter was investigated given the association of T90 with poor CV outcomes in prior studies.^27^ T90 was assessed by median due to its skewed nature and for interpretability; an approach used in prior studies. ^28,29^ Other hypoxia measures were investigated including the mean oxygen saturation (mSaO2) and SaO2 nadir. Natural language processing^30^ was used to obtain documentation of continuous positive airway pressure (CPAP) prescription at the time of the first sleep study.

### Neighborhood Socioeconomic Position

ADI is a census block composite measure of neighborhood disadvantage that uses 17 poverty, education, housing, and employment indicators. It stratifies geographic areas based on socioeconomic disadvantage and is calculated by national rank ranging from 1 to 100 and by state rank from 1 to 10. Higher percentiles, reflect greater neighborhood deprivation and quartiles were constructed from the 25^th^, 50^th^ and 75^th^ percentiles, ADI –Q1 through Q4.^19^ We identified patients’ addresses within the medical record on the date of the sleep study and addresses were geocoded to the census block group and linked to the University of Wisconsin Neighborhood Atlas.^19,31^

### Outcomes

The primary outcome of interest was the incidence of MACE, a composite of cardiovascular outcomes, defined as the first occurrence of cerebrovascular events, heart failure, atrial fibrillation, coronary artery disease, or death. Outcomes were identified by the *International Classification of Diseases, Ninth Revision, and International Classification of Diseases, Tenth Revision* (ICD 9 and ICD 10) codes in clinical and procedural encounters **(codes reported in eTable1 in Supplement).** Death information was extracted from Cleveland Clinic electronic health records, the Ohio Vitals Data (up to 2019), and the National Death Index. We also separately analyzed individual components of MACE, i.e. incident cerebrovascular events, heart failure, atrial fibrillation, coronary artery disease, and death. Identification of the first event of occurrence was after the index date (sleep study date) with a censoring date of the last clinical encounter up to October 18, 2021. The censoring date was chosen based on the most recent upgrade and revision of the sleep study registry.

### Statistical Analysis

Data are presented as mean ± standard deviation (SD) or median [25th, 75th percentiles] for continuous variables and counts (percentages) for categorical variables. Comparisons of variables across ADI quartiles were made using ANOVA with pairwise testing, adjusted for multiple comparisons. To evaluate the association between ADI and the composite outcome of MACE and all-cause mortality, unadjusted and adjusted Cox proportional hazards models were used to estimate hazard ratios (HR) and 95% confidence intervals (CI) for each quartile of ADI compared to the reference lowest quartile (ADI-Q1). These models were used to determine the hazard risk of the composite outcome of either an initial MACE event or death, right censored to a common date, and adjusted for covariates. Covariates included age, sex, BMI (kg/m^2^), race, cardiovascular medications, smoking status (current or former vs never), individual comorbidities comprising the Elixhauser score, the total Elixhauser score, and OSA-related measures including AHI ≥30 events/hour and T90 dichotomized at the median. AHI cut-off of ≥30 events/hour was chosen given the strong and consistent association of severe OSA with CVD incidence and morbidity.^32–34^ The proportional odds assumption was checked for all models. In addition, the statistical interactions of ADI and OSA-related measures were analyzed. We used Cox proportional hazard models to determine the hazard of the combined outcome (initial MACE or death) for each OSA-related measure, and the proportional hazard method of Fine and Gray^35^ (sub-distribution method) to determine the hazard for MACE (or death) in the presence of death (or MACE) as a competing risk. To evaluate the linear relationship between objective OSA-related measures and ADI, we used general linear models adjusted for age, BMI, gender, payor, and race. Outcomes were transformed (e.g. log, natural log, square root function, etc.) as needed to improve model fit. Results are presented from adjusted models.

Secondary analyses included sensitivity analysis excluding patients on prescribed positive airway pressure (PAP) after the sleep study. Given that hypopnea scoring (3% vs 4% oxygen desaturation) may differentially classify OSA severity,^36,37^ a stratified analysis of hypopnea definition, i.e. 3% desaturation or EEG microarousal compared with 4% desaturation was conducted. Lastly analysis of incidence of individual components of MACE by ADI was performed. All statistical analyses were performed based on an overall significance level of 0.05, using SAS software (version 9.4, Cary, NC).

## Results

Our final analytic sample was comprised of 72,443 adults **(Figure S1).** Areas of least and greatest deprivation were seen in both urban and rural areas of north east Ohio counties **(Figure 1)** The mean age was 50.4 ± 14.2 years and included 35,875 (49.5%) women and 36,559 (50.5%) men of whom 52,868 (73.0%) were non-Hispanic White, 13,326 (18.4%) non-Hispanic Black and 2,443 (3.4%) were Hispanic or Latino. Demographics and clinical characteristics are summarized in **Table 1**. Compared to residents living in areas of least deprivation (ADI-Q1), those in areas of greatest deprivation (ADI- Q4) were more likely to be women: 7,393 (40.1%) vs 10,432 (59.9%), p<.001, younger: 52.3 ± 14.0 vs 48.9 ± 13.9, p<.001, of black race: 898 (4.9%) vs 8,529 (49.0%), p<.001 with a higher BMI: 31.9 ± 7.3 vs 37.3 ± 9.2, p<.001) respectively, and to have more comorbidities (increased Elixhauser Comorbidity Index Score, 1.1 ± 1.2 vs 1.5 ± 1.4, p<.001).

**Figure 1.**
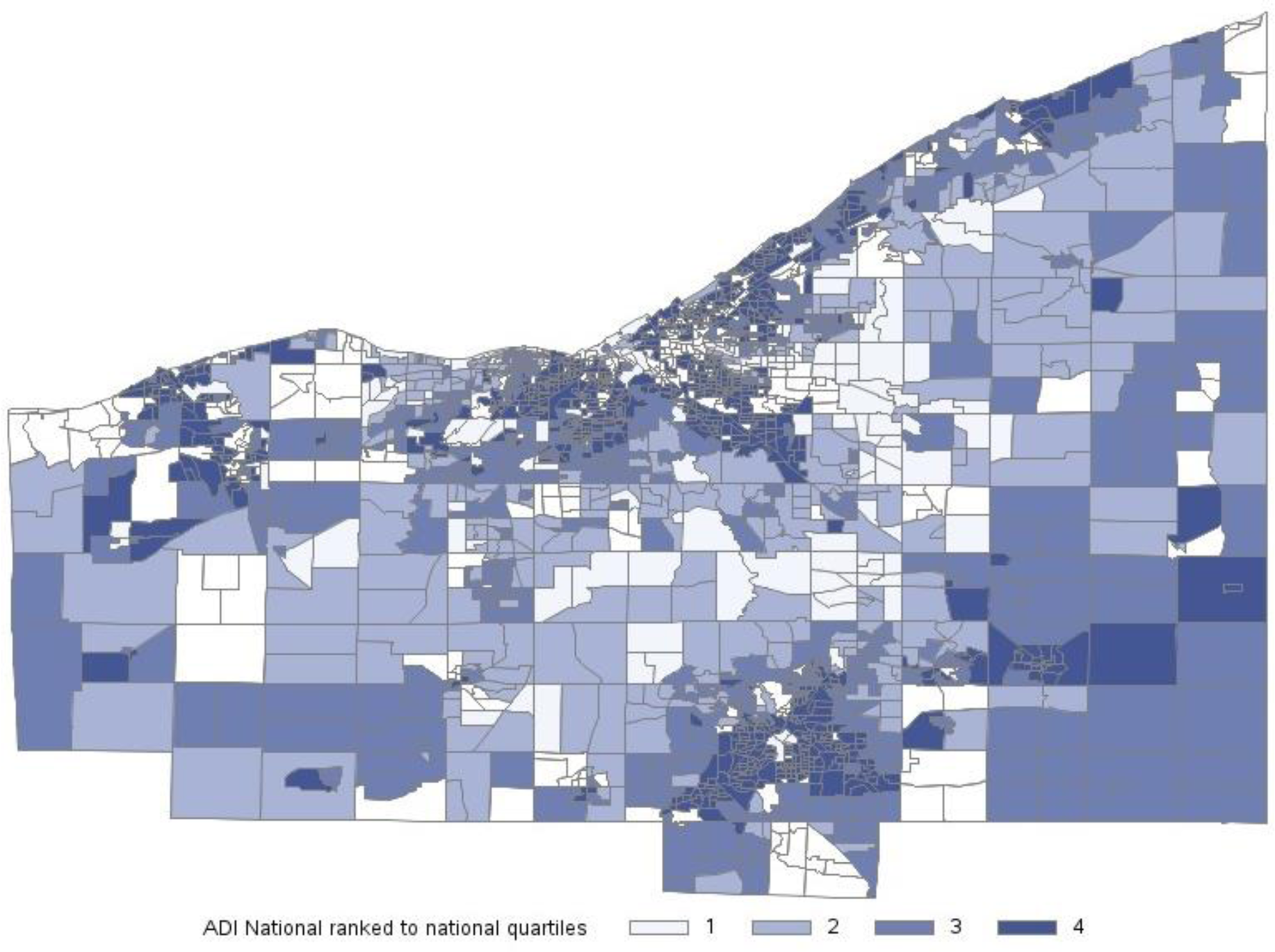
Areas of Deprivation Index Distribution in Quartiles Across Northeast Ohio Counties

**Table 1.** Demographic, Comorbidities, and Sleep Study elements, all study eligible patients (n=72,443)

| Factor | Total<br>(N=72,443) | Q1<br>(N=18,438) | Q2<br>(N=18,484) | Q3<br>(N=18,103) | Q4<br>(N=17,418) | p-value |
| --- | --- | --- | --- | --- | --- | --- |
| **Basic Demographics** |  |  |  |  |  |  |
| Age (years) | 50.4 ± 14.2 | 52.3 ± 14.0<sup>2,3,4</sup> | 51.4 ± 14.4<sup>1,3,4</sup> | 49.1 ± 14.2<sup>1,2</sup> | 48.9 ± 13.9<sup>1,2</sup> | <.001<sup>a</sup> |
| Sex* |  |  |  |  |  | <.001<sup>c</sup> |
| Female | 35,875 (49.5) | 7,393 (40.1)<sup>2,3,4</sup> | 8,629 (46.7)<sup>1,3,4</sup> | 9,421 (52.0)<sup>1,2,4</sup> | 10,432 (59.9)<sup>1,2,3</sup> |  |
| Male | 36,559 (50.5) | 11,042 (59.9) | 9,853 (53.3) | 8,679 (48.0) | 6,985 (40.1) |  |
| Race/Ethnicity |  |  |  |  |  | <.001<sup>c</sup> |
| Non-Hispanic White | 52,868 (73.0) | 16,116 (87.4) | 15,920 (86.1) | 13,873 (76.6) | 6,959 (40.0) |  |
| American Indian or Alaska Native | 71 (0.10) | 18 (0.10)<sup>2,3,4</sup> | 17 (0.09)<sup>1,3,4</sup> | 22 (0.12)<sup>1,2,4</sup> | 14 (0.08)<sup>1,2,3</sup> |  |
| Asian/Pacific Islander | 823 (1.1) | 385 (2.1) | 171 (0.93) | 191 (1.06) | 76 (0.44) |  |
| Non-Hispanic Black | 13,326 (18.4) | 898 (4.9) | 1,246 (6.7) | 2,653 (14.7) | 8,529 (49.0) |  |
| Hispanic (any race) | 2,443 (3.4) | 267 (1.4) | 343 (1.9) | 559 (3.1) | 1,274 (7.3) |  |
| Multiracial | 791 (1.09) | 186 (1.01) | 199 (1.08) | 230 (1.3) | 176 (1.01) |  |
| Other or not stated | 2,121 (2.9) | 568 (3.1) | 588 (3.2) | 575 (3.2) | 390 (2.2) |  |
| Body mass index, kg/m<sup>2</sup>, mean ± SD | 34.4 ± 8.5 | 31.9 ± 7.3<sup>2,3,4</sup> | 33.7 ± 8.0<sup>1,3,4</sup> | 35.1 ± 8.6<sup>1,2,4</sup> | 37.3 ± 9.2<sup>1,2,3</sup> | <.001<sup>a</sup> |
| Primary payor |  |  |  |  |  | <.001<sup>c</sup> |
| Private | 38,669 (53.4) | 11,386 (61.8)<sup>2,3,4</sup> | 10,580 (57.2)<sup>1,3,4</sup> | 9,889 (54.6)<sup>1,2,4</sup> | 6,814 (39.1)<sup>1,2,3</sup> |  |
| Medicaid | 8,031 (11.1) | 641 (3.5) | 1,264 (6.8) | 2,055 (11.4) | 4,071 (23.4) |  |
| Medicare | 20,926 (28.9) | 5,220 (28.3) | 5,488 (29.7) | 4,862 (26.9) | 5,356 (30.7) |  |
| Self-pay | 3,510 (4.8) | 937 (5.1) | 849 (4.6) | 943 (5.2) | 781 (4.5) |  |
| Other/ Not reported | 1,307 (1.8) | 254 (1.4) | 303 (1.6) | 354 (2.0) | 396 (2.3) |  |
| ADI State Rank, mean | 4.5 ± 3.0 | 1.2 ± 0.38<sup>2,3,4</sup> | 2.7 ± 0.71<sup>1,3,4</sup> | 5.4 ± 0.99<sup>1,2,4</sup> | 8.8 ± 0.99<sup>1,2,3</sup> | <.001<sup>a</sup> |
| ADI State Rank, median | 4.0 [2.0, 7.0] | 1.00 [1.00, 1.00]<sup>2,3,4</sup> | 3.0 [2.0, 3.0]<sup>1,3,4</sup> | 5.0 [5.0, 6.0]<sup>1,2,4</sup> | 9.0 [8.0, 10.0]<sup>1,2,3</sup> | <.001<sup>b</sup> |
| Epworth Sleepiness Scale* | 9.3 ± 5.2 | 8.8 ± 4.9<sup>2,3,4</sup> | 9.2 ± 5.1<sup>1,3,4</sup> | 9.5 ± 5.2<sup>1,2,4</sup> | 9.9 ± 5.5<sup>1,2,3</sup> | <.001<sup>a</sup> |
| **Comorbidities** |  |  |  |  |  |  |
| Obesity | 19,304 (26.6) | 3,551 (19.3)<sup>2,3,4</sup> | 4,476 (24.2)<sup>1,3,4</sup> | 5,199 (28.7)<sup>1,2,4</sup> | 6,078 (34.9)<sup>1,2,3</sup> | <.001<sup>c</sup> |
| Diabetes | 9,557 (13.2) | 1,763 (9.6)<sup>2,3,4</sup> | 2,253 (12.2)<sup>1,3,4</sup> | 2,425 (13.4)<sup>1,2,4</sup> | 3,116 (17.9)<sup>1,2,3</sup> | <.001<sup>c</sup> |
| Hypertension | 21,989 (30.4) | 5,180 (28.1)<sup>4</sup> | 5,417 (29.3)<sup>4</sup> | 5,266 (29.1)<sup>4</sup> | 6,126 (35.2)<sup>1,2,3</sup> | <.001<sup>c</sup> |
| Heart Failure | 317 (0.44) | 83 (0.45) | 78 (0.42) | 92 (0.51) | 64 (0.37) | 0.24<sup>c</sup> |
| Any cancers | 4,346 (6.0) | 1,257 (6.8)<sup>3,4</sup> | 1,218 (6.6)<sup>3,4</sup> | 985 (5.4)<sup>1,2</sup> | 886 (5.1)<sup>1,2</sup> | <.001<sup>c</sup> |
| Asthma | 14,031 (19.4) | 2,769 (15.0)<sup>2,3,4</sup> | 3,093 (16.7)<sup>1,3,4</sup> | 3,596 (19.9)<sup>1,2,4</sup> | 4,573 (26.3)<sup>1,2,3</sup> | <.001<sup>c</sup> |
| COPD | 4,764 (6.6) | 746 (4.0)<sup>2,3,4</sup> | 1,034 (5.6)<sup>1,3,4</sup> | 1,263 (7.0)<sup>1,2,4</sup> | 1,721 (9.9)<sup>1,2,3</sup> | <.001<sup>c</sup> |
| Smoking Status |  |  |  |  |  | <.001<sup>c</sup> |
| Never | 37,725 (52.1) | 10,890 (59.1)<sup>2,3,4</sup> | 9,761 (52.8)<sup>1,3,4</sup> | 9,067 (50.1)<sup>1,2,4</sup> | 8,007 (46.0)<sup>1,2,3</sup> |  |
| Former | 22,308 (30.8) | 5,485 (29.7) | 5,885 (31.8) | 5,708 (31.5) | 5,230 (30.0) |  |
| Passive smoker | 512 (0.71) | 123 (0.67) | 127 (0.69) | 146 (0.81) | 116 (0.67) |  |
| Current | 8,768 (12.1) | 1,235 (6.7) | 1,940 (10.5) | 2,390 (13.2) | 3,203 (18.4) |  |
| Unknown | 3,130 (4.3) | 705 (3.8) | 771 (4.2) | 792 (4.4) | 862 (4.9) |  |
| Total Elixhauser Commodity Index | 1.3 ± 1.4 | 1.1 ± 1.2<sup>2,3,4</sup> | 1.2 ± 1.3<sup>1,3,4</sup> | 1.3 ± 1.4<sup>1,2,4</sup> | 1.5 ± 1.4<sup>1,2,3</sup> | <.001<sup>a</sup> |
| Total Elixhauser Commodity Index | 1.0 [0.0, 2.0] | 1.0 [0.0, 2.0]<sup>2,3,4</sup> | 1.0 [0.0, 2.0]<sup>1,3,4</sup> | 1.0 [0.0, 2.0]<sup>1,2,4</sup> | 1.0 [0.0, 2.0]<sup>1,2,3</sup> | <.001<sup>e</sup> |
| Total Elixhauser comorbidities |  |  |  |  |  | <.001<sup>c</sup> |
| 0 – 5 | 71,908 (99.3) | 18,360 (99.6)<sup>2,3,4</sup> | 18,364 (99.4)<sup>1,4</sup> | 17,970 (99.3)<sup>1,4</sup> | 17,214 (98.8)<sup>1,2,3</sup> |  |
| >5 | 535 (0.74) | 78 (0.42) | 120 (0.65) | 133 (0.73) | 204 (1.2) |  |
| **Medications** |  |  |  |  |  |  |
| Anti-Arrhythmics | 279 (0.39) | 73 (0.40) | 81 (0.44) | 77 (0.43) | 48 (0.28) | 0.054<sup>c</sup> |
| Beta Blockers | 11,124 (15.4) | 2,565 (13.9)<sup>2,3,4</sup> | 2,936 (15.9)<sup>1</sup> | 2,724 (15.0)<sup>1,4</sup> | 2,899 (16.6)<sup>1,3</sup> | <.001<sup>c</sup> |
| ARBs | 6,532 (9.0) | 1,653 (9.0) | 1,663 (9.0) | 1,547 (8.5)<sup>4</sup> | 1,669 (9.6)<sup>3</sup> | 0.008<sup>c</sup> |
| ACE-I | 11,187 (15.4) | 2,639 (14.3)<sup>2,4</sup> | 2,837 (15.3)<sup>1,4</sup> | 2,744 (15.2)<sup>4</sup> | 2,967 (17.0)<sup>1,2,3</sup> | <.001<sup>c</sup> |
| Aspirin | 7,031 (9.7) | 1,928 (10.5)<sup>2,3</sup> | 1,707 (9.2)<sup>1,3,4</sup> | 1,512 (8.4)<sup>1,2,4</sup> | 1,884 (10.8)<sup>2,3</sup> | <.001<sup>c</sup> |
| Digoxin | 0 (0.00) | 0 (0.00) | 0 (0.00) | 0 (0.00) | 0 (0.00) |  |
| Statins | 17,582 (24.3) | 4,944 (26.8)<sup>2,3,4</sup> | 4,629 (25.0)<sup>1,3,4</sup> | 3,952 (21.8)<sup>1,2,4</sup> | 4,057 (23.3)<sup>1,2,3</sup> | <.001<sup>c</sup> |
| Nitrates | 1,444 (2.0) | 300 (1.6)<sup>2,4</sup> | 381 (2.1)<sup>1</sup> | 333 (1.8)<sup>4</sup> | 430 (2.5)<sup>1,3</sup> | <.001<sup>c</sup> |
| P2Y12 Receptor Blocker | 1,102 (1.5) | 260 (1.4) | 313 (1.7)<sup>3</sup> | 232 (1.3)<sup>2,4</sup> | 297 (1.7)<sup>3</sup> | 0.001<sup>c</sup> |
| **Sleep Measures** |  |  |  |  |  |  |
| Sleep study type |  |  |  |  |  | <.001<sup>c</sup> |
| PSG and Split | 52,874 (73.0) | 12,294 (66.7)<sup>2,3,4</sup> | 12,913 (69.9)<sup>1,3,4</sup> | 13,354 (73.8)<sup>1,2,4</sup> | 14,313 (82.2)<sup>1,2,3</sup> |  |
| Type III | 19,569 (27.0) | 6,144 (33.3) | 5,571 (30.1) | 4,749 (26.2) | 3,105 (17.8) |  |
| Hypopnea scoring rule |  |  |  |  |  | <.001<sup>c</sup> |
| 3% | 50,124 (69.2) | 13,942 (75.6) | 13,265 (71.8) | 12,579 (69.5) | 10,338 (59.4) |  |
| 4% | 22,042 (30.4) | 4,422 (24.0) | 5,145 (27.8) | 5,457 (30.1) | 7,018 (40.3) |  |
| Not reported | 277 (0.38) | 74 (0.40)<sup>2,3,4</sup> | 74 (0.40)<sup>1,3,4</sup> | 67 (0.37)<sup>1,2,4</sup> | 62 (0.36)<sup>1,2,3</sup> |  |
| AHI, median [IQR] | 14.3 [5.8, 33.3] | 15.1 [6.2, 33.1]<sup>3,4</sup> | 14.6 [6.0, 33.0]<sup>4</sup> | 13.8 [5.6, 33.3]<sup>1</sup> | 13.5 [5.4, 34.1]<sup>1,2</sup> | <.001<sup>b</sup> |
| % Sleep Time with SaO<sub>2</sub><90% (T90)* | 3.8 [0.40, 23.6] | 4.4 [0.50, 25.6]<sup>3,4</sup> | 4.4 [0.50, 25.9]<sup>3,4</sup> | 3.6 [0.40, 23.1]<sup>1,2,4</sup> | 2.9 [0.40, 19.0]<sup>1,2,3</sup> | <.001<sup>b</sup> |
| Mean O<sub>2</sub> saturation (%)* | 92.9 ± 2.9 | 92.8 ± 2.7<sup>2,4</sup> | 92.7 ± 2.7<sup>1,3,4</sup> | 92.8 ± 2.9<sup>2,4</sup> | 93.1 ± 3.2<sup>1,2,3</sup> | <.001<sup>a</sup> |
| Minimum O<sub>2</sub> saturation (%)* | 83.1 ± 7.9 | 83.5 ± 7.4<sup>3,4</sup> | 83.3 ± 7.6<sup>4</sup> | 83.1 ± 8.0<sup>1,4</sup> | 82.5 ± 8.6<sup>1,2,3</sup> | <.001<sup>a</sup> |

\*Data not available for all 72,443 subjects. Missing values: ADI Quartiles = 597; BMI = 516; Percent sleep time SaO<sub>2</sub> <90% (T90)= 9,059, Gender Description = 9; Mean O<sub>2</sub> saturation(%) = 1,918; Minimum O<sub>2</sub> saturation= 1,959; Epworth Sleepiness Scale = 1,456.
Abbreviations: BMI; Body mass index; Q: Quartiles; SD: Standard Deviation; ADI: Area of Deprivation Index; COPD: Chronic Obstructive Pulmonary Disease; ARBs: Angiotensin Receptor Blockers; ACE-I: Angiotensin Converting Enzyme Inhibitors (ACEI); AHI: Apnea Hypopnea Index; PSG: polysomnogram; T90 (percentage of sleep time spent with SaO<sub>2</sub><90%); IQR: Interquartile Range.
Statistics presented as Median [P25, P75], N (column %).
p-values: b=Kruskal-Wallis test, c=Pearson's chi-square test, d=Fisher's Exact test, e. Friedman non-parametric ANOVA
<sup>1</sup>: Significantly different from 1st quartile
<sup>2</sup>: Significantly different from 2nd quartile
<sup>3</sup>: Significantly different from 3rd quartile
<sup>4</sup>: Significantly different from 4th quartile
Post-hoc pairwise comparisons were done using Bonferroni adjustment.

### Association of Neighborhood Disadvantage with MACE and Death

The median follow-up was 4.39 [IQR,1.76-7.92] years. At the end of the study period, 8,547 (11.8%) patients experienced a primary composite endpoint (MACE or death) with a higher relative incidence observed among patients living in areas of greatest deprivation (ADI-Q4) compared to patients living in any other area (ADI-Q1-Q3) (p<.001) **(Figure 2).** After adjustment for potential confounders, the primary MACE or death endpoint was 28% higher for individuals living in areas with greatest deprivation (ADI-Q4) compared to areas of lowest deprivation (ADI-Q1) (HR, 1.28[95%CI, 1.20,-1.38]; p<.001) **(Table 2.).** For individuals living in areas of highest deprivation, the risk of death (in 2.3%) with a competing risk of a MACE event was 79% greater than for those living in any other quartile after adjustment for covariates (adjusted-HR, 1.79 [95% CI, 1.51- 2.12]; p<.001) **(Figure 2).**

**Figure 2.**
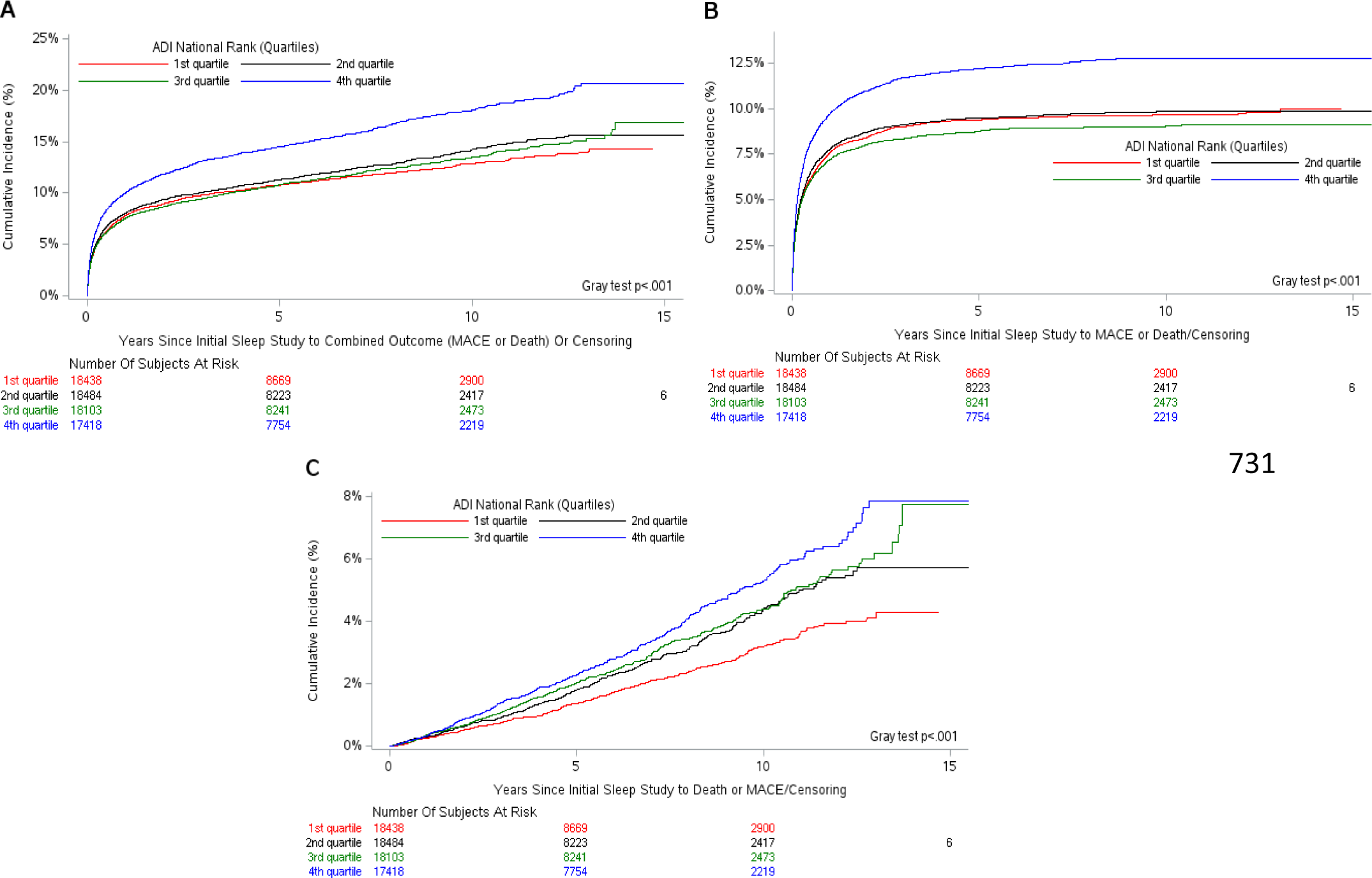
**Cumulative Incidence Function Plots of Outcomes in the Presence of Competing Risks**

**Table 2.** Association of ADI with Primary Outcome MACE or Death in Univariate and Multivariable Analysis using Cox Proportional Hazard Regression (n=72,443) Elixhauser score and Elixhauser score.

| Factor | Univariate Analysis |  | Multivariable Regression |  |
| --- | --- | --- | --- | --- |
|  | HR 95% CI | p value | HR 95% CI | p value |
| Area Deprivation Index, National Rank quartiles (ref: 1st quartile) |  | Overall <.001 |  | Overall <.001 |
| - 2nd quartile | 1.07 (1.01, 1.14) | 0.026 | 1.02 (0.96, 1.09) | 0.518 |
| - 3rd quartile | 1.02 (0.96, 1.08) | 0.582 | 1.02 (0.96, 1.10) | 0.498 |
| - 4th quartile | 1.40 (1.32, 1.48) | <.001 | 1.28 (1.20, 1.38) | <.001 |

Abbreviations: ADI, Area Deprivation Index; MACE, Major Adverse Cardiovascular Events; HR: Hazards ratio; CI: Confidence Interval. Multivariable Analysis was adjusted for age; sex; race; body mass index; T90 above median; AHI > 30 events/hour; cardiac medications including: anti-arrhythmic, beta blockers, aspirin, digoxin, statins, nitrates, P2Y12 receptor blockers, angiotensin receptor blocker, angiotensin converting enzyme inhibitors blockers; smoking status, comorbidities part of the Elixhauser score and Elixhauser score.

Conversely, for patients living in areas of least deprivation and most resources, the risk of MACE with death as a competing risk was 17% higher compared to those living in greater deprivation (adjusted-HR,1.17 [95% CI,1.09, 1.27]; p<.001) **(Figure 2).**

### Linear Relationship of Sleep Disordered Breathing Measures and Neighborhood Deprivation

We observed a significant linear increase between hypoxia measures with increasing area deprivation (in ADI quartiles). In patients who underwent PSG studies, increased T90 (p<.001, decreased mSaO2(p<.001), and decreased SaO2 nadir (p<.001) were associated with increasing ADI. In patients who underwent type III sleep studies, greater ADI (higher quartiles) was associated with decreased mSaO2(p<.001) and decreased SaO2 nadir (p<.001). Likewise, higher AHI was significantly associated with increasing ADI (p=0.003) in patients who underwent PSG; however, no linear relationship was observed between AHI and ADI quartiles among type III sleep study groups (p=0.082).

*Interaction of Sleep Disordered Breathing and Neighborhood Disadvantage on MACE and Death* There was no significant interaction between AHI with ADI on the risk of MACE in the presence of competing risk of death (p=0.586) and on the risk of death in the presence of competing risk of MACE (p=0.614). However, there were significant statistical interactions between T90 with ADI on the risk of MACE in the presence of competing risk of death (p=0.002) and death in the presence of competing risk of MACE (p=0.005). For those living in ADI-Q1 (lowest area deprivation), increased degree of sleep-related hypoxia defined by T90 conferred a 37% increased risk of MACE (adjusted-HR, 1.37[95% CI:1.23 - 1.53]). Among individuals living in ADI-Q4, increased T90 was associated with a 26% increased risk of MACE: adjusted-HR: 1.26 (95% CI:1.14 - 1.38). For those individuals living in ADI Q2 and Q3, T90 was associated with a respective 56% and 51% increased risk of death (adjusted-HR, 1.56[95%CI:1.23-1.96]; adjusted-HR, 1.51[95%CI,1.21- 1.88]), respectively **(Figure 3).**

**Figure 3.**
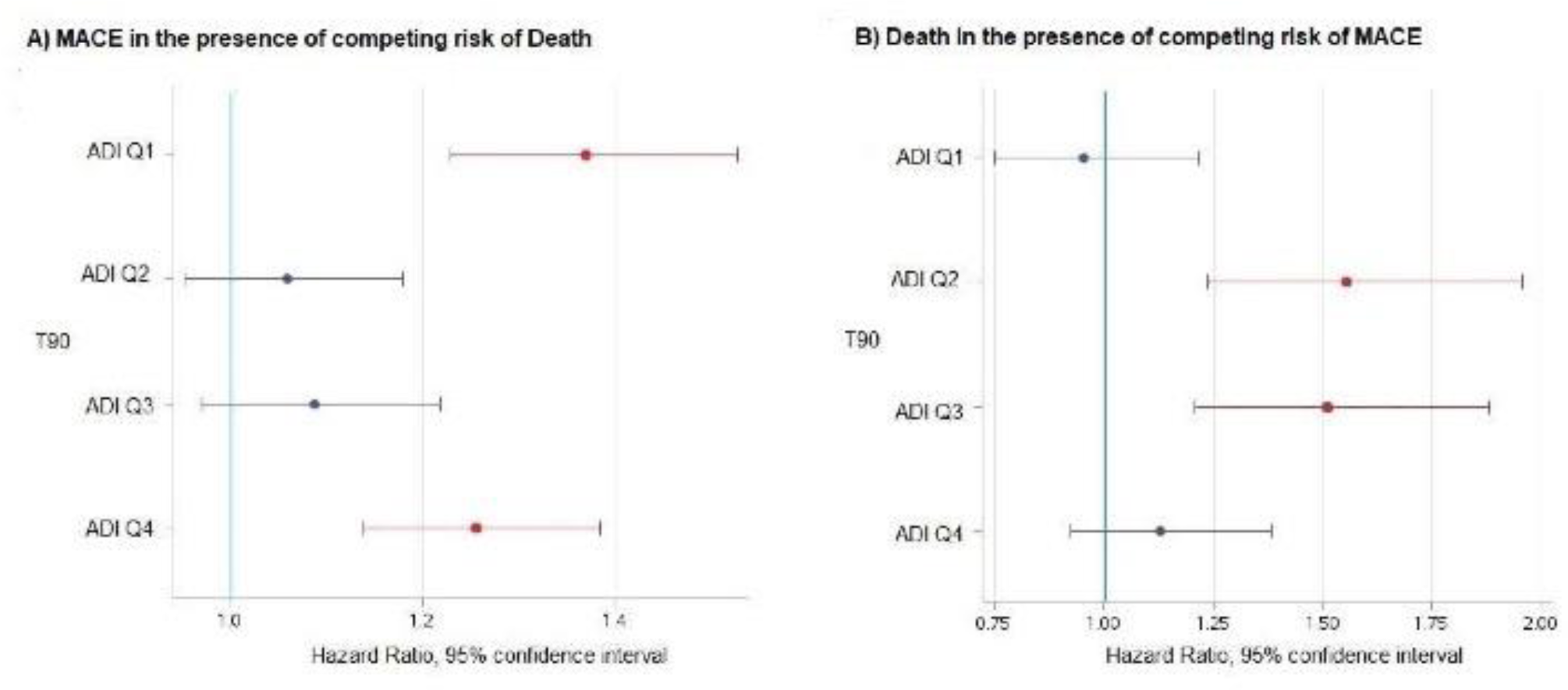
**Effects of ADI and T90 (Interaction) on Adjusted Hazard Ratios and 95% Confidence Limits for MACE and Death Outcomes in the Presence of Competing Risks, T90 and ADI Quartiles**

### Incidence of Individual Components of MACE by Degree of Neighborhood Disadvantage

The cumulative incidence of cerebrovascular events, coronary artery disease, atrial fibrillation, and heart failure was 18.3%, 11.6%, 35.7%, and 37.7% respectively. There was a greater relative incidence seen among patients living in higher areas of deprivation compared to individuals living in low areas of deprivation (p<.001) in each component of MACE except for atrial fibrillation. A higher relative incidence of atrial fibrillation was observed among individuals living in areas of less deprivation compared to individuals living in high areas of deprivation (p<.001) **(Figure 4).**

**Figure 4.**
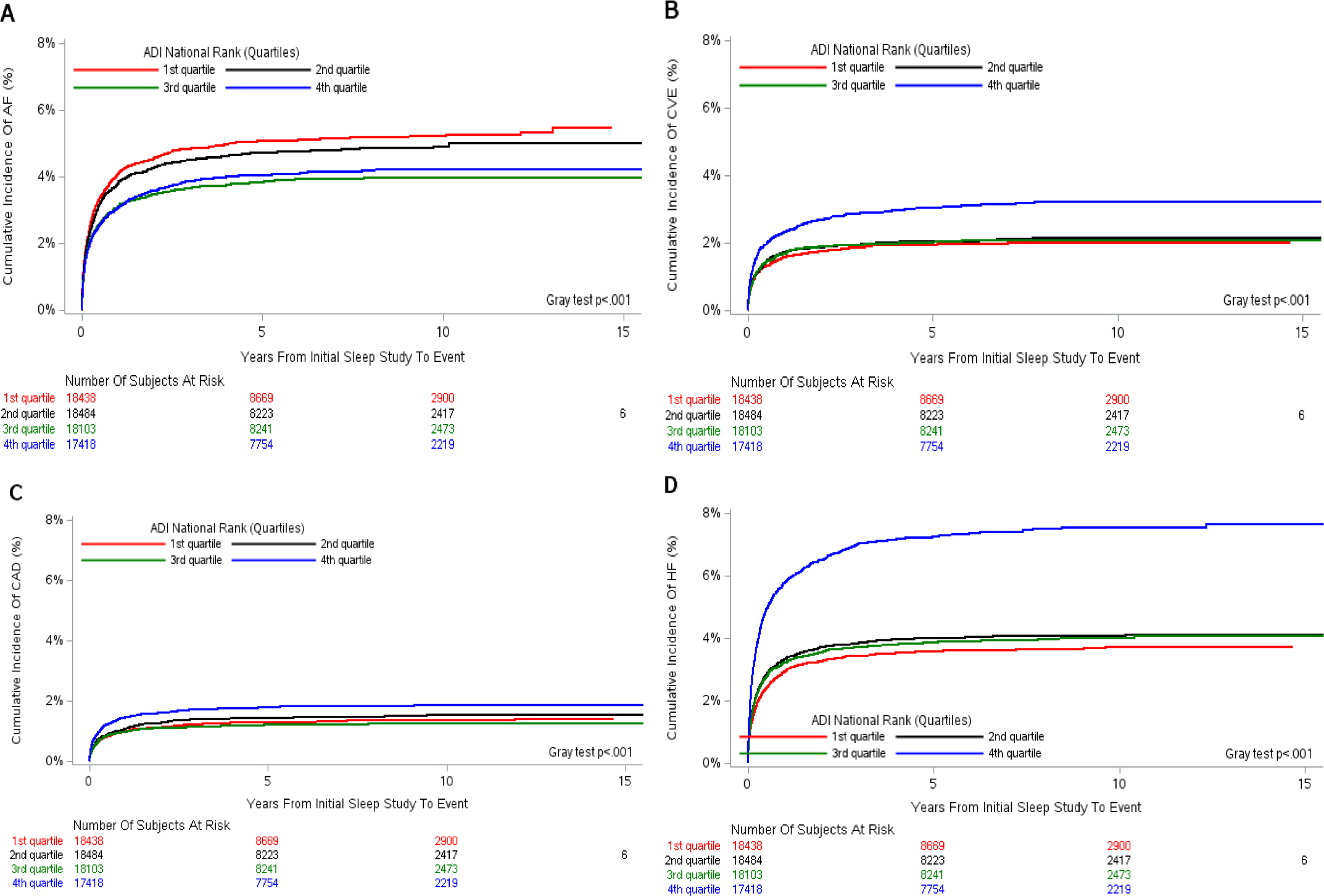
Cumulative Incidence Function Plots for Individual End Points of MACE by ADI Quartile

### Secondary Analysis

After including only 45879 (63%) patients without PAP therapy after the initial sleep study, the results of the analyses of ADI and the primary endpoint of MACE or death were similar to the main results **(Table S3).** Results were also similar for the statistical interaction of T90 and ADI on the risk of MACE in the presence of competing risk of death (p=0.040) and death in the presence of competing risk of MACE (p=0.012) **(Table S4).** In those without PAP therapy, for patients living in areas of lowest deprivation (ADI-Q1), sleep-related hypoxia defined by T90 conferred a 43% increased risk of MACE and an 18% and 27% increased risk of MACE for those living in greatest areas of deprivation (Q3 and Q4) (adjusted-HR, 1.43 [95% CI, 1.26 - 1.62]; adjusted-HR, 1.18 [95% CI:1.04 -1.34]; adjusted-HR, 1.27 [95% CI: 1.14-1.42]). Among those living in ADI-Q2 increased T90 was not associated with an increased risk of MACE (adjusted-HR, 1.13 [95% CI, 0.998 - 1.27] When death was considered, results were consistent with primary results. We also took into consideration the hypopnea definition data which was available in 72,166 (99%) patients, of which 50,124 (69.5%) were scored using the 3% hypopnea definition and 22,042 (30.5%) used the 4% hypopneas definition. Similar to primary findings, in sleep studies using the 3% hypopnea definition, results were similar to the full cohort **(Table S5 and S6).** However, when 4% hypopnea scoring was used, even though the primary MACE or death endpoint was similar to primary results, there was no statistically significant interaction between T90 and ADI **(Table S5 and S7).**

## Discussion

### Principal Findings

We identified that greater versus lower levels of neighborhood deprivation measured by the ADI national rank were associated with increased risk for the composite of MACE and death. Similarly, the degree of sleep-related hypoxia was greater among individuals living in neighborhoods of higher deprivation and this association was more pronounced than the association observed with OSA severity defined by AHI. Moreover, a significant statistical interaction was observed between sleep-related hypoxia with ADI in relation to MACE and death risk. Individuals with increased sleep-related hypoxia living in both extremes of area deprivation (Q4 and Q1) had an increased risk for MACE, whereas individuals with increased sleep-related hypoxia living in moderate areas of deprivation (Q2- Q3) had an increased risk for death. These findings are timely, particularly given the knowledge gaps recently identified in a multi-institute NIH workshop report underscoring the need to better understand the link between sleep health disparities and poor health outcomes to inform targetable inverventions.^38^

### Neighborhood Socioeconomic Disadvantage and Cardiovascular Outcomes

A robust body of literature suggests that neighborhood socioeconomic factors are associated with poor cardiovascular outcomes and CV risk factors for the general population. However, to our knowledge, this is the first study to investigate the association of neighborhood levels of deprivation as it relates to major adverse cardiovascular events in adult patients from a large OSA referral sample phenotyped with overnight sleep studies. Our results thereby highlight the impact of existing neighborhood-related socioeconomic disparities in CVD outcomes among individuals with highly prevalent comorbidities such as OSA. Previous studies have shown that markers of increased neighborhood deprivation such as the social vulnerability index (SVI) and ADI are associated with increased risk for CV risk factors, premature CV mortality, and cardiac readmissions. ^21,39,40^ Moreover, at the county level, United States counties with greater degree of ADI and socioeconomic deprivation are associated with increased premature CV mortality.^41^ However, none of these studies have focused on patients with OSA nor accounted for potential confounders that could have biased the results such as medications, smoking, and obesity, the latter in particular representing a well-recognized risk factor for CVD and mortality.^42–45^ Moreover, SVI measures the social risk of populations and considers age, race, and ethnicity as variables, confounding the overall social risk assessment. On the other hand, ADI allows the ability to exclusively characterize different levels of socioeconomic deprivation from specific areas without the confounding influence of race and ethnicity within the measure. Additionally, and similar to other studies, we observed an increased incidence of CVD among patients living in areas of greater deprivation except for AF. One potential explanation for this observation is the association of age, SEP, in the incidence AF as previous research has found an increased lifetime risk of AF among individuals with higher SEP.^46^

### Neighborhood Socioeconomic Disadvantage and OSA

Our findings suggest that sleep-related hypoxia-- more so than the degree of OSA characterized by frequency of apneas and hypopneas-- is associated with socioeconomic disadvantage. Furthermore, the degree of sleep-related hypoxia increases with greater areas of deprivation which may be explained by different determinants at multiple levels of influence. At the individual level, this association may be explained by the increased comorbid cardiopulmonary conditions, smoking history, or the presence of individual socioeconomic factors in OSA. At the community level, it is highly possible that neighborhood-related environmental factors dictated by racial residential segregation or social cohesion may represent an important factor in these differences. It is known that socially disadvantaged neighborhoods are more exposed to environmental hazards, such as increased air pollution as well as increased light and noise at night which can disturb sleep. Along these lines, increased air pollution is associated with OSA and a reduction in oxygen saturation, ^47–49^ hence serving as a potential explanation for the increased degree of sleep-related hypoxia among individuals living in greater areas of disadvantage.

### Association of OSA and Neighborhood Socioeconomic Disadvantage as it related to MACE and Death

We observe that greater levels of sleep-related hypoxia among individuals living in both extreme levels of socioeconomic deprivation increase the risk of MACE; a finding not observed among those with moderate deprivation. Although understanding factors associated with these results requires further investigation, this observation may be explained by the differences in lifestyle and dietary choices across quartiles, environmental exposures, and lack of concordance between individual and community-level social risks. An increased prevalence of social determinants of health (SDOH) at the individual level across quartiles has been reported, including in quartiles of lowest deprivation in primary care patients. Therefore, differences in the type and prevalence of SDOH seen at the individual level may have a different influence on health outcomes independently of neighborhood-related SDOH. Further studies are needed to investigate SDOH at multiple levels of influence, i.e., individual and community, to better understand the mechanisms that underlie health disparities across ADI quartiles.

In our secondary analysis, while no statistically significant interaction was observed between sleep-related hypoxia and ADI when the cohort was stratified by 4% hypopnea scoring rule, results remained the same when the 3% hypopnea rule was considered. These findings may be explained by either smaller sample size of the 4% hypopnea rule subgroup or possibly point towards the known inequities of using the 4% hypopnea rule in OSA severity.^36,46^

## Limitations

This study has limitations. As common to retrospective studies, our study is susceptible to referral and selection biases. Generalizability of findings can be extrapolated to a clinical referral sample and Ohio-based population. Although we attempted to address influence of underlying lung disease by smoking history, we did not have granular level data on packs per day of exposure nor lung function testing to more rigorously account for pulmonary-specific contributions to sleep-related hypoxia.

Finally, our institution is a quaternary care center and although we included only those with residence in Northeast Ohio, it is possible that patients may have had follow-up outside of our hospital system. Even though this may have resulted in potential underestimation of initial MACE or death, such misclassification would be expected to bias findings towards the null.

## Conclusions

In this study, increased neighborhood socioeconomic disadvantage contributes to an increased risk of MACE and death among patients with increased sleep-related hypoxia. Our findings implicate sleep-related hypoxia as an important target to address disparities in OSA contributing to inequities in CV outcomes. Future research should evaluate specific neighborhood-related factors i.e., noise, light and air pollution, advertisements promoting tobacco and alcohol usage, lack of safe places to exercise which increases the risk of obesity, which may be important determinants of sleep-related hypoxia in OSA, CV health and associated disparities.

## Non-standard Abbreviations and Acronyms

ADI: Area Deprivation
Index AHI: Apnea Hypopnea Index
CPAP: Continuous Positive Pressure
CV: Cardiovascular
OSA: Obstructive Sleep Apnea
PSG: Polysomnogram
SEP: Socioeconomic Position
STARTLIT: Sleep Signals, Testing, and Reports Linked to Patients Traits
SaO2: Oxygen Saturation
mSaO2: Mean Oxygen Saturation
T90: Percentage of sleep time spent<90% of oxygen saturation

## Acknowledgments

### Author contributions

Authors were involved in the conception and design of the study, data collection, data analysis and interpretation. All authors provided critical revision of the article and approved the final version of the manuscript. The manuscript was reviewed and edited by all the authors. All authors made the decision to submit the manuscript for publication and assume responsibility for the accuracy and completeness of the analyses and for the fidelity of this report to the study methods.

Conception and design: RM, CPO

Data collection, analysis, and interpretation: RM, CPO, DB

Drafting and revision of the manuscript for important intellectual content: All authors Statistical Analysis: DB, RM, CPO

CPO, RM and DB had full access to all the data in the study and takes responsibility for the integrity of the data and the accuracy of the data analysis and include this in the Acknowledgment section of the manuscript.

### Sources of Founding

Neuroscience Transformative Research Resource Development Award and the Center of Population Health Research at Cleveland Clinic.

### Supplemental Material

**Supplemental Methods.** Sleep Testing, Registry, and Respiratory Event Scoring

**Table S1.** Cardiac Medications and Comorbidities

**Table S2.** Diagnosis and Procedures Codes

Figure S1. Identification of Eligible Patients of the Analytic Sample

### Secondary Analysis

**Table S3.** Association of Area Deprivation Index with Primary Outcome MACE or Death Univariate and Multivariable Analysis without PAP Therapy after Sleep Study

**Table S4.** Interactions between Obstructive Sleep Apnea Measures with Area Deprivation Index as it related to MACE in those without Positive Airway Pressure therapy

**Table S5.** Association of Area Deprivation Index with Primary outcome MACE or Death Univariate and Multivariable Stratified by Hypopnea rule 3% and 4%

**Table S6.** Interactions Between Obstructive Sleep Apnea Measures with Area Deprivation Index as It Relates to MACE in those with 3% Hypopnea Rule

**Table S7.** Interactions Between Obstructive Sleep Apnea Measures with Area Deprivation Index as It Relates to MACE in those with 4% Hypopnea Rule

Reference #26#

## Data Availability

All data related to the manuscript is available upon request.

---

Figure 1.

*Northeast Ohio Counties by Area Deprivation Index.*

Abbreviations: ADI, area deprivation index.

Figure 2.

*Panel A) Probability of MACE or Death end points, (ADI Q4 vs ADI Q1) HR, 1.28 (95%CI, 1.20-1.38) p<.001; Panel B) Probability of MACE versus competing risk of Death, (ADI Q4 vs ADI Q1) HR, 1.17 (95% CI, 1.09-1.27) p<.001; Panel C) Probability of Death versus competing risk of MACE (ADI Q4 vs ADI Q1) HR, 1.79 (95% CI, 1.51-2.12) p<.001).*

Abbreviations: ADI, area deprivation index; HR, hazard ratio.

Figure 3.

*A) Risk of MACE in the presence of competing risk of death: For ADI Q1 and T90, HR, 1.37 (95% CI, 1.23-1.53); For ADI Q4 and T90, HR, 1.26 (95% CI, 1.14-1.38); and B) Risk of Death in the presence of competing risk of MACE: For ADI Q2 and T90, HR, 1.56 (95% CI, 1.23-1.96); For ADI Q3, HR, 1.51 (95% CI, 1.21-1.88)*.

Abbreviations: T90, percentage of sleep time spent with SaO2<90%; ADI, area deprivation index.

Figure 4.

*For each of the 4 individual outcomes (except of death), any patient with previous history of each individual outcomes prior sleep study date was excluded for risk assessment only for that outcome.* Abbreviations: ADI, area deprivation index; AF, atrial fibrillation; CVE, cerebrovascular events; CAD, coronary artery disease; HF, Heart Failure.

